# SARS-CoV-2 neutralizing antibodies; longevity, breadth, and evasion by emerging viral variants

**DOI:** 10.1101/2020.12.19.20248567

**Authors:** Fiona Tea, Alberto Ospina Stella, Anupriya Aggarwal, David Ross Darley, Deepti Pilli, Daniele Vitale, Vera Merheb, Fiona X. Z. Lee, Philip Cunningham, Gregory J. Walker, David A. Brown, William D. Rawlinson, Sonia R. Isaacs, Vennila Mathivanan, Markus Hoffman, Stefan Pöhlmann, Dominic E. Dwyer, Rebeca Rockett, Vitali Sintchenko, Veronica C. Hoad, David O. Irving, Gregory J. Dore, Iain B. Gosbell, Anthony D. Kelleher, Gail V. Matthews, Fabienne Brilot, Stuart G Turville

## Abstract

The SARS-CoV-2 antibody neutralization response and its evasion by emerging viral variants are unknown. Antibody immunoreactivity against SARS-CoV-2 antigens and Spike variants, inhibition of Spike-driven virus-cell fusion, and infectious SARS-CoV-2 neutralization were characterized in 807 serial samples from 233 RT-PCR-confirmed COVID-19 individuals with detailed demographics and followed up to seven months. A broad and sustained polyantigenic immunoreactivity against SARS-CoV-2 Spike, Membrane, and Nucleocapsid proteins, along with high viral neutralization were associated with COVID-19 severity. A subgroup of ‘high responders’ maintained high neutralizing responses over time, representing ideal convalescent plasma therapy donors. Antibodies generated against SARS-CoV-2 during the first COVID-19 wave had reduced immunoreactivity and neutralization potency to emerging Spike variants. Accurate monitoring of SARS-CoV-2 antibody responses would be essential for selection of optimal plasma donors and vaccine monitoring and design.

**One Sentence Summary:** Neutralizing antibody responses to SARS-CoV-2 are sustained, associated with COVID19 severity, and evaded by emerging viral variants

## Introduction

Control of the SARS-CoV-2 pandemic relies on population resistance to infection due to a post-infection and vaccination-induced immunity. Current questions relate to the level, breadth, and longevity of generated immunity, and whether mutation of the virus will compromise immunity. Previous studies reported varying results in longitudinal changes of the virus-specific antibody response. Some detected stable antibody titers 4-6 months after diagnosis (*1, 2*), while others reported waning of the antibody response 2-3 months after infection (*3, 4*). Differences in assay sensitivity and antigen targets may account for these discrepancies, with Spike and nucleocapsid being the main antigens investigated. Immunoreactivity to other abundant antigens, such as Membrane or Envelope, are unknown. Neutralization of SARS-CoV-2 has been reported for antibodies that bind to Spike, a large homo-trimeric glycoprotein studded across the viral surface (*5, 6*), whereas Membrane and Envelope proteins, although exposed on the viral surface, remain to be identified as neutralizing antibody targets. Rapid development of neutralizing antibody response to Spike correlates with viral immunity, and individuals who seroconvert may develop a lasting neutralization response (*7*).

The SARS-CoV-2 virus has accumulated many polymorphisms across its genome, especially within the Spike gene (*8*). Shortly after the introduction of SARS-CoV-2 into the human population, many early and dominant amino acid polymorphisms were associated with viral entry fitness, such as D614G (*9, 10*). However, the pressure of the neutralising antibody response might select for escape mutations in Spike that limit post-infectious immunity or vaccine protection (*11*). One example is the S477N/D614G Spike variant which appeared in Australia during July and August 2020 and was traced to a single event from Australian hotel quarantine (*12*). The S477N/D614G Spike variant currently represents greater than 5% of Spike variants worldwide, 15% in Europe, and 58% in Oceania (*13*).

Using the lessons learned from research of other viral pathogens and neuroimmunological autoantibodies (*14, 15*), we have developed a suite of novel high-content assays that sensitively assess antibody responses against the native oligomeric structure of Spike and its emerging variants (*16*). To measure the neutralizing capacity, we have also developed a Biosafety Level 2 surrogate Spike-driven virus-cell fusion assay that has been cross-validated with a novel high content, machine-scored, Biosafety Level 3 authentic SARS-CoV-2 neutralization assay.

Herein we characterize the longevity, polyantigenic breadth, and neutralization capacity of the SARS-CoV-2 antibody response in individuals and their responses to globally emerging SARS-CoV-2 variants. Using two longitudinal SARS-CoV-2 community- and hospital-based Australian cohorts representative of the broad spectrum of disease severity at acute infection, we showed that the polyantigenic and neutralizing responses to SARS-CoV-2 are sustained, associated with COVID19 severity, and are evaded by emerging viral variants.

This work provides a community snapshot of humoral immunity in those recovering from infection and sheds light on important considerations for vaccine design and selection of donors for convalescent plasma therapy. Additionally, the modular assays used herein can be adapted for novel viral pathogens to respond rapidly to emerging pathogens.

## Results

### SARS-CoV-2 antibody responses are sustained for up to seven months post-infection and are focused on Spike

SARS-CoV-2 antibodies were assessed in RT-PCR-confirmed COVID-19 convalescent adults in two Australian cohorts; ADAPT, a hospital-based cohort of patients recruited during the first and second wave of infection in Australia (n=83 and n=17), and LIFE, a cohort of plasma donors (n=159) (Table 1, Fig. 1A). Antibody immunoreactivity to SARS-CoV-2 antigens, inhibition of virus-cell fusion, live SARS-CoV-2 neutralization, and immunoreactivity to Spike emerging variants were assessed and antibody features were compared with demographic data (Fig. 1A). At first date of collection post-infection, 96% (81/83 ADAPT, median 71 days, mean 74 days, post first PCR positivity) and 98% (152/159 LIFE, median 59 days, mean 61 days) of infected patients were Spike IgG-positive, and 81% (66/83 ADAPT) and 91% (139/152 LIFE) were Spike IgM-positive (Table 2). A broad range of Spike IgG levels was observed. No differences in Spike IgG and IgM levels were observed between females and males, but higher IgG and IgM levels were associated with older age (P<0.0001) (Fig. S1). Detection of convalescent positive serostatus was more sensitive when Spike IgG were detected by live cell flow cytometry compared to Nucleocapsid IgG or Spike IgG using commercial assays (Table 2).

**Table 1.**
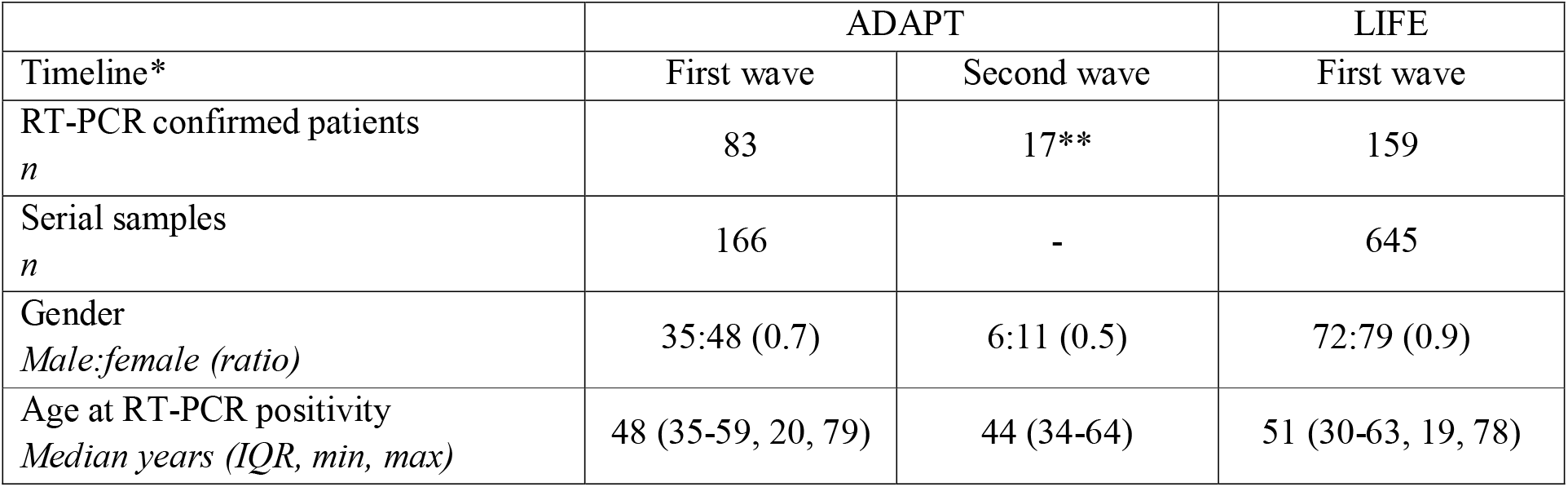
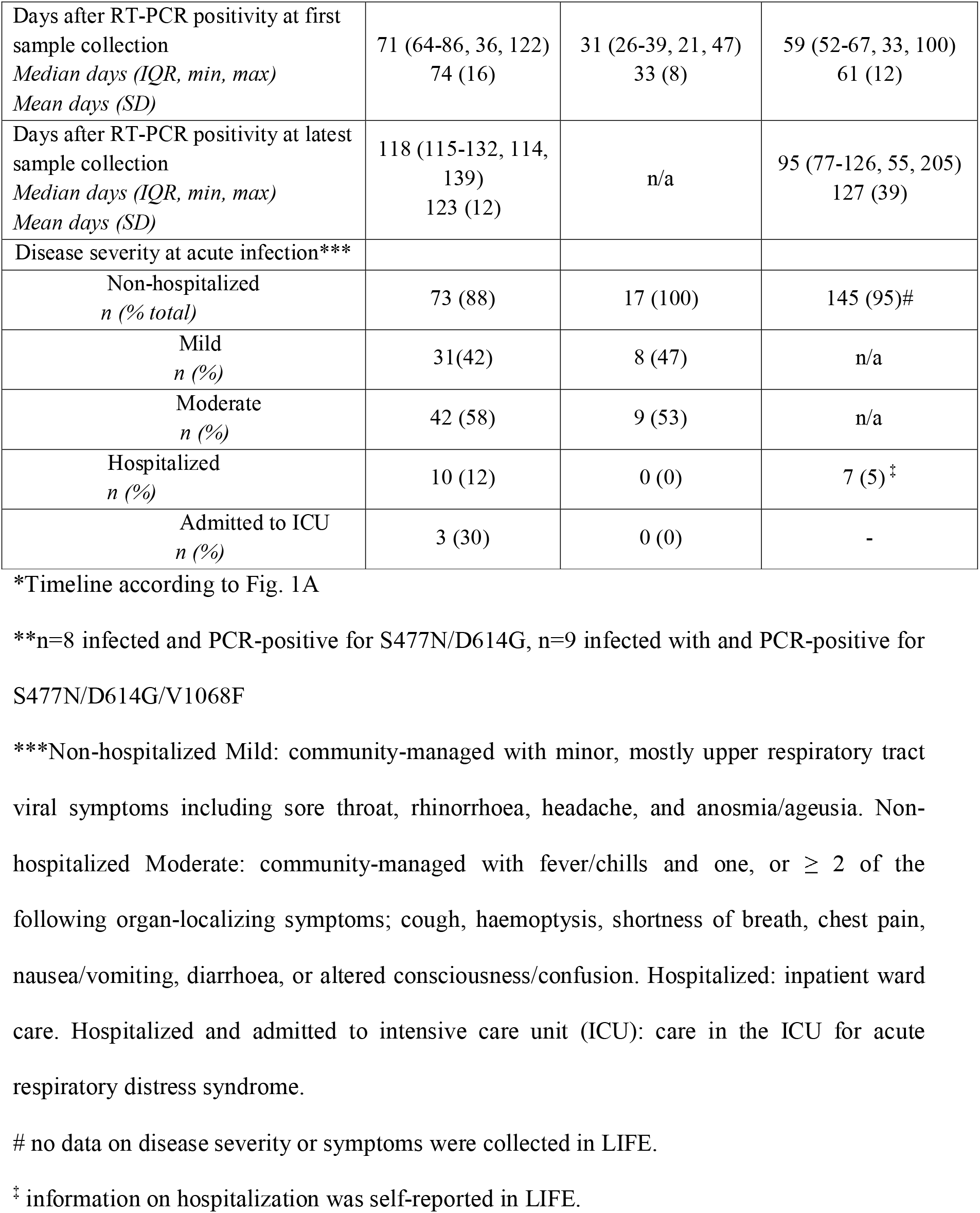
Demographics of the convalescent SARS-CoV-2 ADAPT and LIFE cohorts.

|  | ADAPT |  | LIFE |
| --- | --- | --- | --- |
| Timeline* | First wave | Second wave | First wave |
| RT-PCR confirmed patients<br>*n* | 83 | 17** | 159 |
| Serial samples<br>*n* | 166 | - | 645 |
| Gender<br>*Male:female (ratio)* | 35:48 (0.7) | 6:11 (0.5) | 72:79 (0.9) |
| Age at RT-PCR positivity<br>*Median years (IQR, min, max)* | 48 (35-59, 20, 79) | 44 (34-64) | 51 (30-63, 19, 78) |
| Days after RT-PCR positivity at first sample collection<br>*Median days (IQR, min, max)*<br>*Mean days (SD)* | 71 (64-86, 36, 122)<br>74 (16) | 31 (26-39, 21, 47)<br>33 (8) | 59 (52-67, 33, 100)<br>61 (12) |
| Days after RT-PCR positivity at latest sample collection<br>*Median days (IQR, min, max)*<br>*Mean days (SD)* | 118 (115-132, 114, 139)<br>123 (12) | n/a | 95 (77-126, 55, 205)<br>127 (39) |
| Disease severity at acute infection*** |  |  |  |
| Non-hospitalized<br>*n (%) total* | 73 (88) | 17 (100) | 145 (95)# |
| Mild<br>*n (%)* | 31(42) | 8 (47) | n/a |
| Moderate<br>*n (%)* | 42 (58) | 9 (53) | n/a |
| Hospitalized<br>*n (%)* | 10 (12) | 0 (0) | 7 (5) ‡ |
| Admitted to ICU<br>*n (%)* | 3 (30) | 0 (0) | - |

\*Timeline according to Fig. 1A
\*\*n=8 infected and PCR-positive for S477N/D614G, n=9 infected with and PCR-positive for S477N/D614G/V1068F
\*\*\*Non-hospitalized Mild: community-managed with minor, mostly upper respiratory tract viral symptoms including sore throat, rhinorrhoea, headache, and anosmia/ageusia. Non-hospitalized Moderate: community-managed with fever/chills and one, or $\geq 2$ of the following organ-localizing symptoms; cough, haemoptysis, shortness of breath, chest pain, nausea/vomiting, diarrhoea, or altered consciousness/confusion. Hospitalized: inpatient ward care. Hospitalized and admitted to intensive care unit (ICU): care in the ICU for acute respiratory distress syndrome.

**Table 2.** Comparison of the sensitivity of SARS-CoV-2 antibody detection assays.

|  | ADAPT<br>n=166 samples |  | LIFE |  |  |  |
| --- | --- | --- | --- | --- | --- | --- |
|  |  |  | All samples<br>n=645 |  | First and last samples<br>n=302* |  |
|  | Positive<br>samples, n<br>(%) | Sensitivity<br>% (95%<br>CI) | Positive<br>samples, n<br>(%) | Sensitivity<br>% (95%<br>CI) | Positive<br>samples, n<br>(%) | Sensitivity<br>% (95%<br>CI) |
| **Spike IgG** |  |  |  |  |  |  |
| Flow cytometry assay | 162 (98) | 98<br>(94-99) | 645 | n/a | 302 | n/a |
| Euroimmun | 121 (73) | 73<br>(65-80) | n/a | n/a | n/a | n/a |
| **Spike IgM** |  |  |  |  |  |  |
| Flow cytometry assay | 127 (77) | 76**<br>(70-83) | 608 (94) | 94<sup>^^§</sup><br>(92-96) | 276 (91) | 91<sup>^^§</sup><br>(87-94) |
| **S1/S2 Spike IgG** |  |  |  |  |  |  |
| DiaSorin Liason SARS-CoV-2 S1/S2 IgG assay | 134 (81) | 81<br>(74-86) | n/a | n/a | n/a | n/a |
| **Nucleocapsid IgG** |  |  |  |  |  |  |
| Abbott Architect SARS-CoV-2 assay | 116 (70) | 70<br>(62-77) | 472 (73) | 73<br>(70-77) | 222 (74) | 74<br>(68-78) |
| Euroimmun | 121 (73) | 73<br>(65-80) | n/a | n/a | n/a | n/a |
| **Nucleocapsid/Spike IgG\*\*\*** | n/a | n/a | 577 (89) | 89<br>(87-92) | 271 (90) | 90<br>(86-93) |
| **Membrane IgG** |  |  |  |  |  |  |
| Flow cytometry assay | 87 (52) | 52<br>(45-60) | n/a | n/a | 173 (57) | 57<br>(51-63) |
| **Envelope IgG** | 4 (2) | 2 (0.8-6) | 0 | 0 | 0 | 0 |

\*Only first and last samples of LIFE cohort were tested for Membrane IgG
\*\*Sensitivity is influenced by IgM sero-reversion in 5 ADAPT and 14 LIFE donors
\*\*\*Positivity determined using a two-step clinical diagnostic testing with the Abbott Architect SARS-CoV-2 Nucleocapsid IgG assay, followed by the Euroimmun Spike IgG assay.

**Fig 1.**
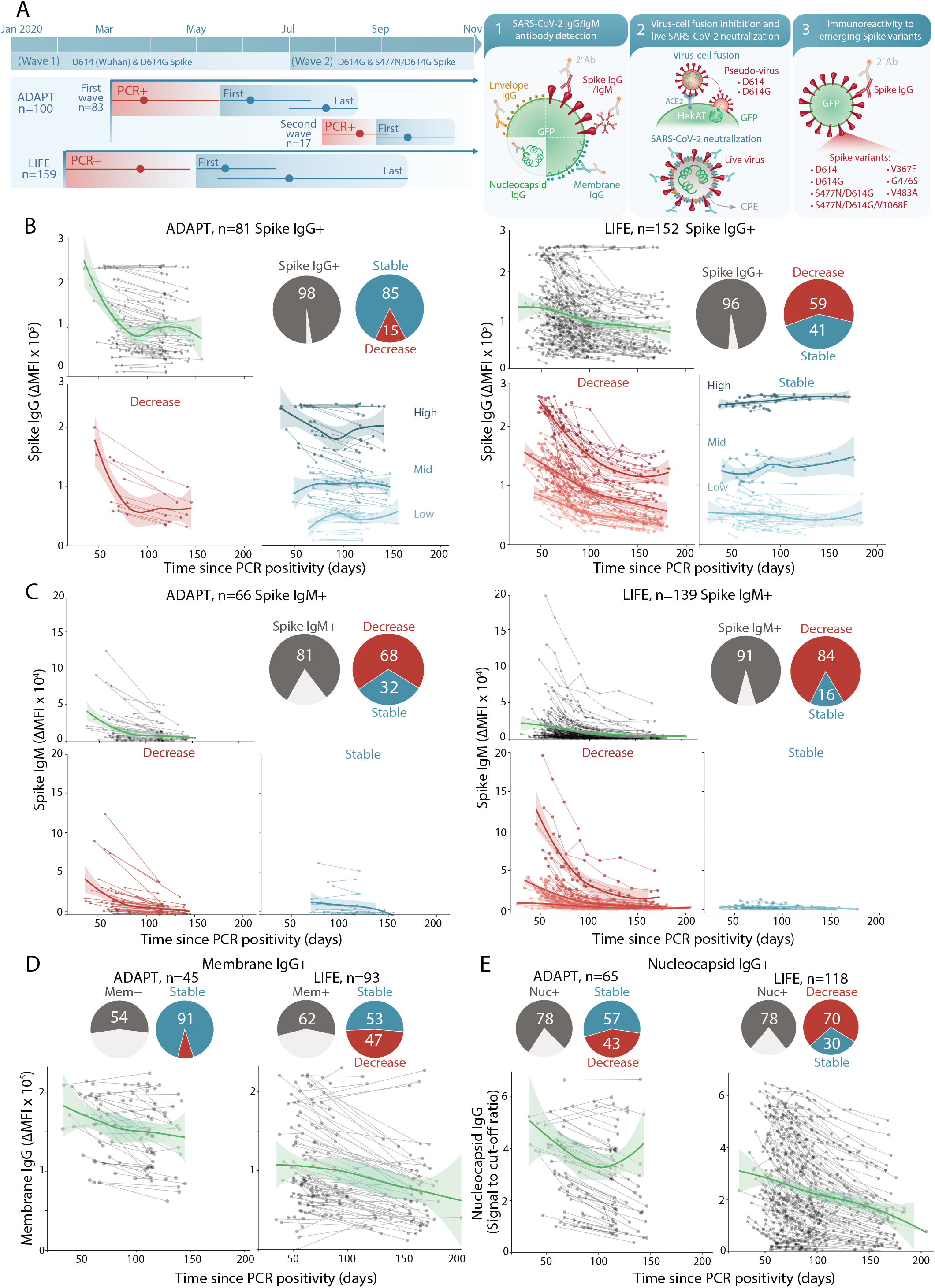
SARS-CoV-2 antibody responses are sustained and are predominantly focused on Spike. (**A**) The first wave of Australian infections were from D614 and D614G Spike, and the S477N/D614G Spike variant emerged during the second wave. Convalescent patient sera from ADAPT (first and second waves) and LIFE (first wave) were examined for SARS-CoV-2 antibodies. Mean time and range of PCR-positivity (red), and dates of first and last sample collection (blue) are shown. Seropositive patients with at least three weeks between first and last samples were examined over time. **(B)** 96-98% (grey) of patients were Spike IgG+. Most ADAPT patients had stable levels overtime, whereas most of LIFE Spike IgG levels decreased. No patients seroreverted. **(C)** 81-91% (grey) were Spike IgM+, most had decreasing levels over time and Spike IgM+ individuals started with and maintained low IgM levels. **(D)** 54-57% (grey) of sera were Membrane IgG+, and most ADAPT had stable levels, whereas a larger proportion of LIFE had decreasing levels. **(E)** 78% of sera were Nucleocapsid IgG+, most were stable in ADAPT, whereas most decreased in LIFE. Loess curves with 95% confidence intervals are shown.

The longevity of antibody responses was assessed in 807 Spike IgG-positive serial samples from 233 individuals (n=162 ADAPT, n=645 LIFE), spanning up to 205 days post-PCR positivity (Fig. 1A and Table 1). There was a range of Spike IgG titers at first collection date, and among all Spike IgG-positive individuals, no individual seroreverted, even up to 205 days post-PCR positivity. The majority of ADAPT patients had stable IgG responses (85%), whereas most LIFE donors exhibited decreased IgG over time (59%), where a decrease was defined as >30% change from first collected sample (Fig. 1B) (*14*). A two-phase decay in those with decreasing responses characterized by an initial high rate of decay followed by stabilization, and the breakpoint between the two phases was estimated at 85 days post-PCR positivity (Fig. 1B). The level at which Spike IgG stabilized was dependent on intial antibody response. High Spike IgG levels decayed to mid-level reactivity and mid-low level reactivity to low level (Fig. 1B). In Spike IgM-positive patients, the majority had decreased IgM levels over time (68% ADAPT; 84% LIFE), in which levels initially decreased and then stabilised at lower levels, but did not serorevert up to 205 days (Fig. 1C). Only five ADAPT (6%) and fourteen LIFE (9%) individuals sero-reverted for Spike IgM at median 146 days post-PCR positivity (Fig. 1C). The breakpoint between the two phases of IgM decay was at 93 days post-PCR positivity.

The polyantigenic breadth of Spike IgG-positive individuals against the virus was examined by detecting IgG targeting the SARS-CoV-2 Membrane, Envelope, and Nucleocapsid proteins (Table 2). 54% (45/83 ADAPT) and 57% (87/152 LIFE) individuals harboured IgG targeting the SARS-CoV-2 Membrane protein, whereas 78% had antibody targeting the Nucleocapsid protein (65/83 ADAPT, 118/152 LIFE) (Fig. 1D and 1E). Antibody titers toward the Membrane protein remained stable over the period of observation in most individuals (91%, 41/45 ADAPT; 95%, 83/87, LIFE), whereas responses toward the Nucleocapsid protein differed between ADAPT and LIFE, and were reminiscent of the Spike IgG response, i.e. mostly stable in ADAPT, and mostly decreased in LIFE over time (Fig. 1D and 1E). Across both cohorts, reactivity to the Envelope protein was very limited with only two ADAPT patients (2%) positive for Envelope IgG (Table 2). Antibody responses to SARS-CoV-2 were highly focused on Spike, followed by the Nucleocapsid and Membrane proteins. Individuals with higher Spike IgG had also high levels of Nucleocapsid and Membrane IgG (Fig. S1).

The overall decay of SARS-CoV-2 antibodies between both cohorts behaved similarly for Spike IgM, but not for Spike IgG, Membrane IgG, and Nucleocapsid IgG, with LIFE donors exhibiting a higher proportion of decreased profiles (Fig. 1). The first collected sample in ADAPT started later post-PCR positivity, and the time duration between paired samples was shorter than for LIFE samples, therefore some ADAPT patients may have been captured during the 2^nd^, more stable, phase (Fig. 1 and Table 1). Furthermore, few ADAPT patients underwent plasmapheresis, whereas all LIFE donors underwent plasmapheresis as part of convalescent plasma donations (median 6 donations, IQR 3-9, max 14). However, donors with more than 10 donations (n=30) had decay profiles similar to the whole cohort, in which donors stabilized at mid-low level, and none of these highly recurrent donors became seronegative (Fig. S2).

### Neutralization of SARS-CoV-2 is correlated with Spike antibody levels and is maintained over time

The neutralization capacity of these individual responses was assessed on a Spike-driven virus-cell fusion assay and a whole-virus neutralization assay (Fig. 1A). Most sera were capable of inhibiting virus-cell fusion (82%, 68/83 ADAPT; 68%, 104/152 LIFE) and mediating viral neutralization (88%, 73/83 ADAPT; 94%, 143/152 LIFE) (Fig. 2A, Table 2). In both cohorts, the virus-cell fusion assay was more stringent than the SARS-CoV-2 neutralization assay as a proportion of individual sera with lower titers in the SARS-CoV-2 neutralization assay were negative in the virus-cell fusion assay (7%, 6/83 ADAPT; 27%, 41/152 LIFE), and most individuals had higher titers in the neutralization assay (Fig. 2A). To understand the discrepancy between both viral assays, live SARS-CoV-2 viral particles were enumerated and directly compared to Spike-pseudotyped lentiviral particles. Cell permeable RNA-specific staining of live virions detected viral particles that were Nucleocapsid-positive (Fig. 2B). The particle to transduction ratios from the fusion assay were 1.03×10^5^, consistent with the low specific infectivity of lentiviruses such as HIV-1(*17*). In contrast, the SARS-CoV-2 particle to infectivity ranged from 58 (HekAT14) to 578 (VeroE6), consistent with the ratio reported for influenza virus (*18*). However, the absolute viral particle number was 74-fold higher in Spike-pseudotyped particle preparation (1.64×10^8^ particles per ml) compared to authentic SARS-CoV-2 (2.22×10^6^ particles per ml). Thus, the specific infectivity of SARS-CoV-2 was higher than that of Spike-expressing lentiviral particles, which may account for the higher sensitivity of the SARS-CoV-2-based neutralization assay.

**Fig 2.**
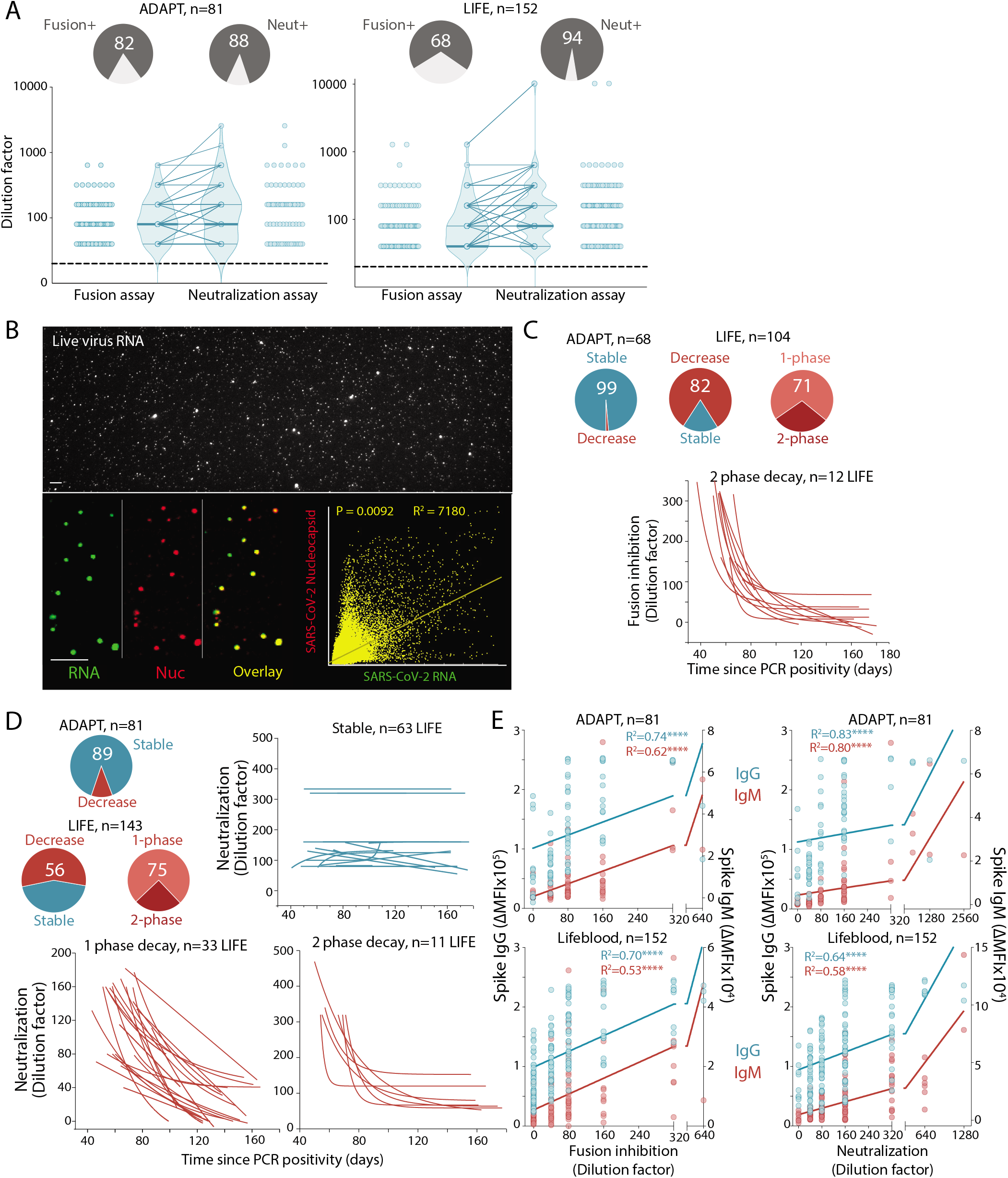
Viral neutralization and inhibition of viral-cell fusion are strongly correlated to Spike antibody titers, and sustained overtime. **(A)** 68-82% of convalescent sera inhibited virus-cell fusion, whereas 88-94% sera neutralized live authentic SARS-CoV-2. **(B)** ∼75% of virus particles were SARS-CoV-2 Nucleocapsid- and RNA-positive (overlay, yellow) **(C)** All but one ADAPT patient had stable responses over time, whereas most LIFE donors (82%) had a decreased virus-cell fusion over time, with the majority (71%) exhibiting a single-phase decay. **(D)** In sera capable of viral neutralization, most ADAPT sera were stable (89%), whereas most LIFE sera (56%) had a decreased score over time, with the majority (75%) exhibiting a single-phase decay. Serum curves unable to be fitted were classified as undetermined. **(E)** Spike IgG and IgM levels were correlated to inhibition of virus-cell fusion and neutralization scores. R^2^ values are shown and ∗ indicates significance.

Most ADAPT patients had stable virus-cell fusion inhibition (99%) and neutralization (89%) titers over time (Fig. 2C and 2D). Most of LIFE donors had decreased virus-cell fusion inhibition (82%) and neutralization (56%) capacity over time, and the majority exhibited a single-phase decay in both assays (Fig. 2C and 2D). The greater number of samples per LIFE donor enabled finer characterization of the decay profile in 34 donors in the virus-cell fusion and 44 donors in the neutralization assay). Most donors had a single-phase decay, whilst a two-phase decay was observed in those with >1:320 titers at first collection. These rapidly dropped and then stabilized over time at 1:80 to 1:160 (28% and 25% of LIFE donors in the virus-cell fusion and neutralization assays, respectively). Individuals with two-phase decay had much higher starting titers than individuals with a single-phase decay (Fig. 2D). In the neutralization assay, LIFE donors with decreased profile had a similar median follow up as the stable profile (∼63 days and 56 days, respectively). In both cohorts, the neutralization and fusion profiles were similar to the Spike IgG profiles, in which ADAPT had more stable responses than LIFE. Indeed, Spike IgG and IgM titers were strongly correlated with virus-cell fusion inhibition and SARS-CoV-2 neutralization (Fig. 2E).

### A broad antigenic repertoire and high neutralization capacity against SARS-CoV-2 is associated with COVID-19 severity

Approximately half of individuals (55% ADAPT and 49% LIFE) had broad polyantigenic immunoreactivity as defined by IgG responses against each of SARS-CoV-2 Spike, Membrane, and Nucleocapsid proteins (Fig. 3A). Around a third of individuals exhibited antibodies against only two proteins (27% and 30%, Nucleocapsid and Spike; 2% and 9% toward Membrane and Spike in ADAPT and LIFE respectively), and a smaller proportion had responses against Spike alone (12 and 17%) (Fig. 3A). Polyantigenic immunoreactivity did not change overtime in most individuals (82%, 15/81 ADAPT, 83%, 41/152 LIFE, data not shown). No individual developed IgG to new antigens at any point of follow up, but instead, lost immunoreactivity to one antigen, either Nucleocapsid or Membrane.

**Fig 3.**
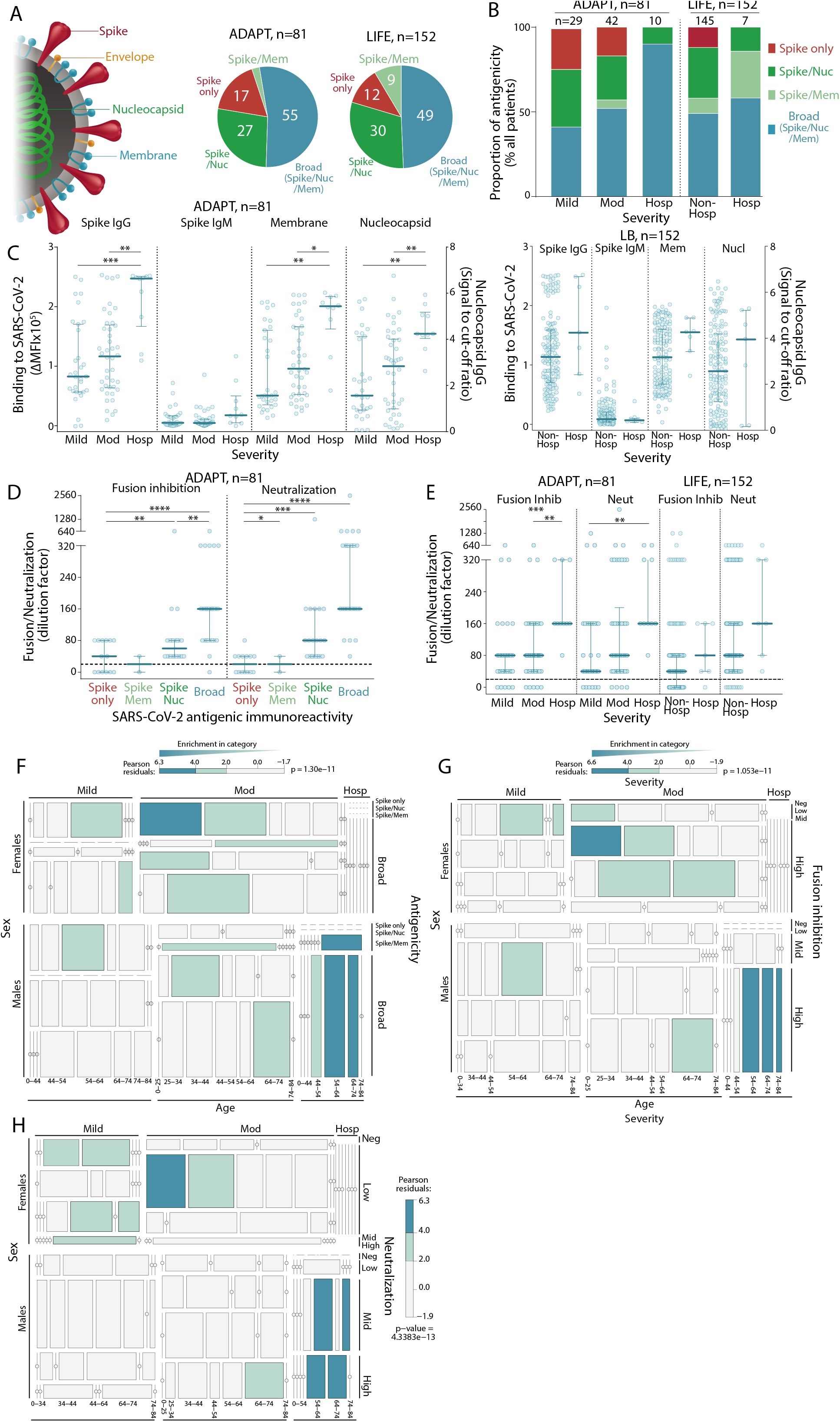
The antibody responses of patients with more severe COVID-19 disease have broader SARS-CoV-2 polyantigenicity. **(A)** ∼ half of patients (49-55%) had broad SARS-CoV-2 antibodies (blue). Some had responses to two antigens (light and dark green), and a few reacted to Spike only (red). **(B)** Hospitalized patients were more likely to have broad SARS-CoV-2 polyantigenic immunoreactivity, whereas patients with only Spike reactivity exhibited mild-moderate symptoms. **(C)** Hospitalized patients exhibited higher Spike IgG, IgM, Membrane IgG, and Nucleocapsid IgG levels. High virus-cell fusion inhibition and neutralization titers were observed in patients with broad polyantigenic immunoreactivity **(D)** and in hospitalized patients **(E)**. Older males were more likely to present with broader polyantigenic immunoreactivity **(F)**, higher virus-cell fusion inhibition **(G)**, and neutralization scores **(H)**. Younger females were more enriched in mild to moderate disease severity, with narrow antigenicity **(F)**, and lower virus-cell fusion inhibition **(G)** and neutralization scores **(H)**.

Patients had broader responses across the spectrum of severity in ADAPT (Fig. 3B). ADAPT and LIFE hospitalized patients with more severe symptoms were more likely to exhibit a broader antibody response to SARS-CoV-2, i.e. polyreactive toward the three antigens (Fig. 3B). Interestingly, two of seven hospitalized LIFE patients who had a short 24 hour hospitalization harboured non-broad responses, and Spike-only responses were exclusively observed in non-hospitalized, mild, and moderate individuals (Fig. 3B). Higher IgG titers against Membrane and Nucleocapsid proteins were also associated with disease severity in both cohorts (Fig. 3C). Patients with broader SARS-CoV-2 responses and higher disease severity had greater viral neutralization and virus-cell fusion inhibition (Fig. 3D, E). This polyreactive, high severity subgroup was populated almost exclusively by older males (Fig. 3F). Similarly, higher neutralization and virus-cell fusion inhibition titers were more enriched in older males with moderate disease and who were hospitalized (Fig. 3G and H).

### High responders with strong and broad SARS-CoV-2 antibody responses are ideal plasma donors

A small subgroup of individuals were “high responders” characterized by high Spike IgG, Spike IgM-positive, broad polyantigenic immunoreactivity (binding to Nucleocapsid, Spike, and Membrane), virus-cell fusion inhibition (>1:160), and neutralization (>1:320). They maintained this high response over time (n=14, 17% ADAPT, n=19, 12% LIFE). High responders were more likely to be male, hospitalized, and were of older age (Fig. 4A). Further characterization was performed on a series of increasingly permissive cell lines, VeroE6, HekAT14, HekAT10, and HekAT24 (Fig. 4B and Fig. S3). Low, i.e. non-high responders, and high responders sera neutralized live SARS-CoV-2 in VeroE6, HekAT14, and HekAT10 cell lines, whereas limited neutralization was observed in the hyper-permissive HekAT24 cell line (Fig. 4B). Using the HekAT24 cell line, two elite responders were identified in LIFE (Fig. 4C), with high Spike IgG and IgM levels, and neutralization titers 30- to 4-fold greater than other individuals (Fig. 4D). Interestingly, elite responders had the highest detectable IgM levels, and early IgM decay coincided with a decrease in neutralization titers, whereas Spike IgG remained stable overtime (Fig. 4E). This association between decreased IgM and neutralization titers was observed in ∼10% of individuals in both cohorts (data not shown).

**Fig 4.**
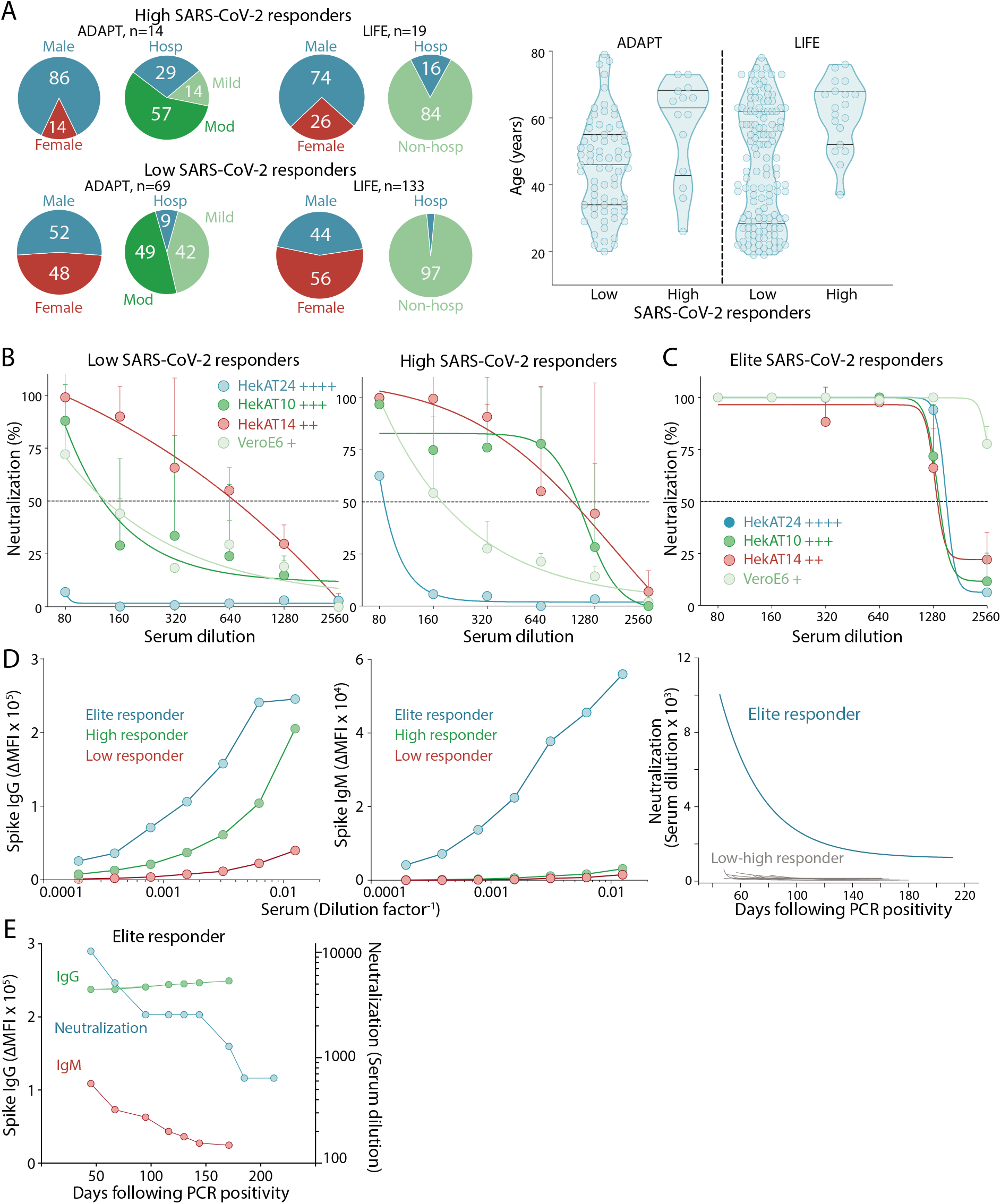
High and elite responders are discriminated with SARS-CoV-2-permissive cells. **(A)** Patients with high and robust SARS-CoV-2 responses were more likely male, hospitalised (left), and of older age (right). **(B)** Low and high responders to SARS-CoV-2 showed limited neutralization in hyperpermissive HekAT24 clonal cells. Permissiveness is indicated by +. **(C)** Only Elite responders showed neutralization in HekAT24 cells. **(D)** Serum titration curves from an Elite responder (blue) showed IgG and IgM levels greater than low (red) and high (green) responders, and incredibly high neutralization titers (≥10,000) that decreased and stabilized at high levels (≥1280). **(E)** The elite donor demonstrated stable high Spike IgG, but the early decrease in viral neutralization was parallel to IgM decline before stabilization (at high titer).

### Spike IgG antibody binding and neutralizing capacity are dependent on Spike mutations in emerging new variants

Numerous Spike polymorphisms have evolved over the course of the pandemic (*11*) with the most attention given to the transmission fitness gain variants, such as D614G (*10, 11*). To test the breadth of the antibody response, Spike IgG immunoreactivity to several Spike variants implicated in the Receptor Binding Domain (RBD) and S1 was assessed (Fig. 1A). Expression of all Spike variants was similar across each transfected cell line used in the flow cytometry antibody assays (Fig. S4A). Compared to the Wuhan-1 D614 variant, most patients had similar binding and were able to recognise the Spike RBD variants G476F, V483A, and V367S (Fig. 5A). However, across both cohorts, there was an overall reduced binding to D614G, a prominent non-RBD S1 variant present during the Australian first wave (Fig. 5A). 65% of ADAPT and 91% of LIFE individuals, infected from the first world-wide wave, generated antibodies that bound broadly to G476F, V483A, V367S, and D614G Spike, whereas 35% of ADAPT and 9% of LIFE had more restricted Spike recognition; i.e. they recognized G476F, V483A, V367S, but had a decreased binding to D641G (Fig. 5A). Immunoreactivity toward all Spike variants was stable overtime in most patients (data not shown).

**Fig 5.**
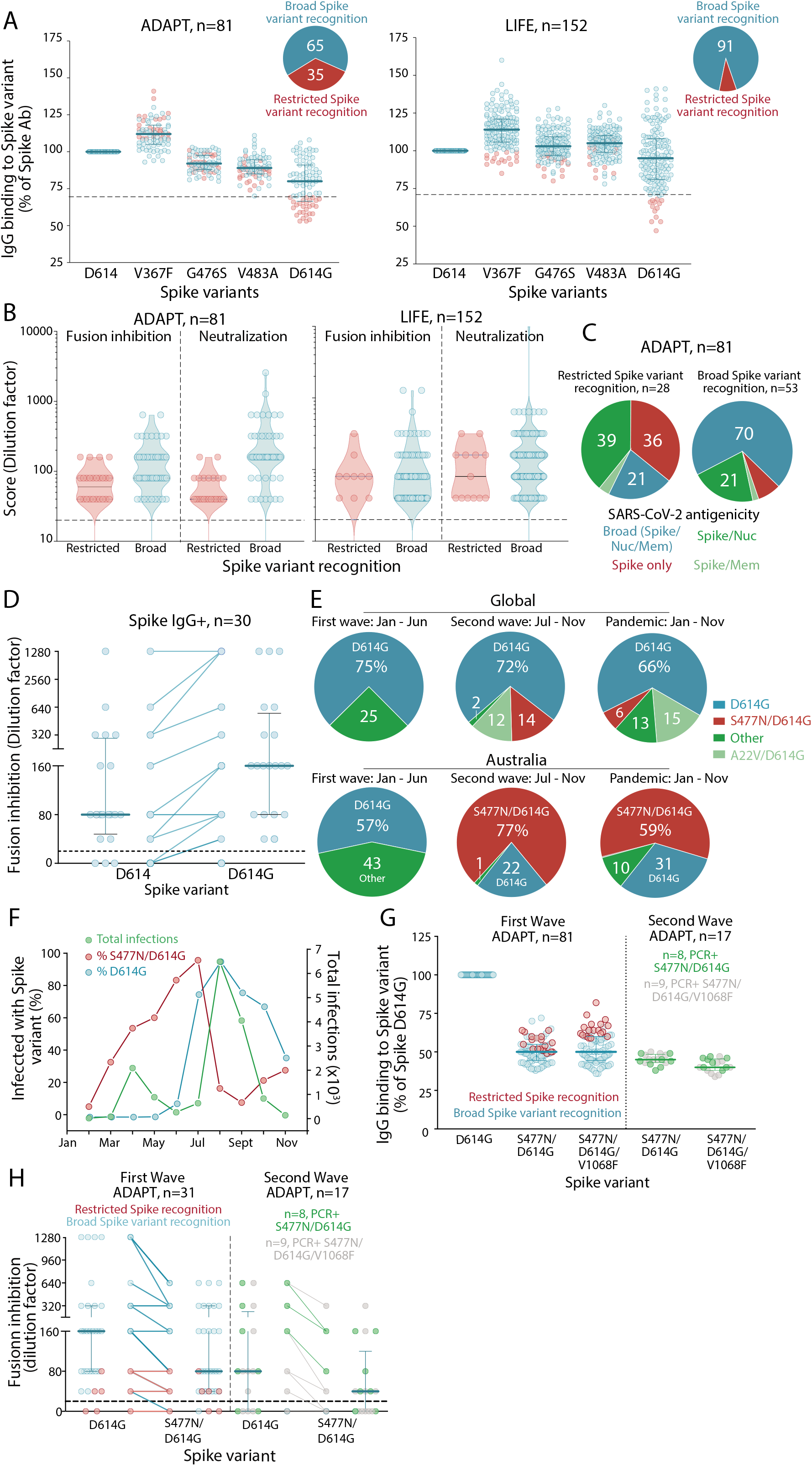
SARS-CoV-2 antibody responses show evasion to emerging Spike variants. **(A)** Most patients had broad recognition of Spike variants (blue), whereas a smaller group had restricted Spike variant recognition and did not have a strong immunoreactivity to D614G Spike (red). Patients with reduced binding to D614G Spike had lower virus-cell fusion and neutralization scores **(B)**, and presented with less broad polyantigenic SARS-CoV-2 recognition **(C). (D)** D614G Spike-binding sera had greater inhibition of D614G Spike-pseudotyped virus-cell fusion. **(E, F)** In Australia, D614G Spike was the predominant variant during the first wave and acquired additional mutations during the second wave (S477N, V1068F). **(G)** All patients had decreased immunoreactivity to S477N/D614G and S477N/D614G/V1068F Spike, while V1068F did not have an additive effect. **(H)** Patients had reduced virus-cell fusion inhibition to the S477N/D614G Spike variant compared to D614G. The level of decreased binding **(G)** and virus-cell fusion inhibition **(H)** was irrespective of the virus that infected patients during the second wave.

Importantly, sera with reduced D614G IgG binding also had lower neutralization and virus-cell fusion inhibition compared to those who recognised D614G Spike (Fig. 5B), suggesting implications for blocking infection in patients who cannot induce robust Spike antibody recognition. Furthermore, patients who bound D614G Spike had broad SARS-CoV-2 polyantigenic immunoreactivity, whereas patients who displayed reduced binding to D614G had more limited antigenic recognition, with 36% recognising Spike only (Fig. 5C). In a D614G virus-cell fusion assay, patients who maintained binding to D614G showed enhanced virus-cell fusion inhibition, compared to when parental Wuhan-1 D614 Spike was used (Fig. 5D). Individuals with lower IgG binding to D614G, i.e. restricted variant recognition had limited D614G Spike virus-cell fusion inhibition, and most (8/11) were unable to prevent Spike fusion (data not shown), emphasizing the need to maintain robust binding to Spike variants for efficient viral neutralization. Patients with restricted Spike variant recognition were not distinguished by age and severity, but were more likely to be female (Fig. S5).

Although D614G Spike remains a predominant variant globally (Fig. 5E), in the second wave of Australian infection between July to September, an isolate with additional polymorphisms, primarily S477N, and in some cases an additional V1068F, was identified. These variants were not detected during the first Australian wave (Fig. 5F), which included the original Wuhan-1 D614 or the D614G variant equally. To assess the antibody binding capacity between original and emerging variants, patients infected by two Spike variants, S477N/D614G and S477N/D614G/V1068F, were recruited during the second wave in Australia (n=17, from the ADAPT cohort, Table 1). All ADAPT patients from the first and second wave had detectable IgG against all Spike variants (Fig. S4B). Compared to the D614G variant, a strong decrease in immunoreactivity to S477N/D614G and S477N/D614G/V1068F was observed in all ADAPT patients from the second wave, whereas the third mutation within the Spike S2 domain V1068F did not have an additive effect (Fig. 5G). This decrease was also observed irrespective of the virus variant that had infected the ADAPT patients (Fig. 5G). Importantly ADAPT patients from the first wave, who had not encountered the new variants, had reduced binding to S477N/D614G and S477N/D614G/V1068F, suggesting a global decrease of immunoreactivity toward both new variants (Fig. 5G). To determine the functional implications of this reduced antibody binding, 48 Spike IgG-positive ADAPT patients (n=31 first wave, n=17 second wave) were assessed for S477N/D614G virus-cell fusion inhibition. Seven patient sera were unable to inhibit virus-cell fusion (Fig. 5H). Compared to D614G Spike, most patients had reduced S477N/D614G Spike virus-cell fusion inhibition (66%, 27/41), and 34% had similar responses (14/41) (Fig. 5H). Interestingly, patients with reduced S477N/D614G Spike virus-cell fusion inhibition had less antibody binding to S477N/D614G Spike than patients with similar fusion inhibition, emphasizing the importance of robust Spike binding for potent viral neutralization.

## Discussion

The current study characterizes the breadth, longevity, and neutralizing capacity of SARS-CoV-2 antibody response in two Australian cohorts, encompassing a wide range of demographics and disease states, up to seven months after COVID-19 diagnosis. We show the development of broad and sustained immunoreactivity against SARS-CoV-2 antigens, and found high titers of Spike-binding and virus-neutralizing antibodies were associated with COVID-19 severity. A group of high responders were identified with high, broad, and sustained neutralizing responses, who may represent ideal donors for convalescent plasma donations. Most importantly, although most patients seroconverted, antibodies generated after early infection displayed a significantly reduced antibody binding and neutralization potency to emerging evasive variants. Our data has important implications on hyperimmune therapy, monoclonal antibody treatments, and vaccine development strategies against emerging viral variants.

The longevity of the immune response against SARS-CoV-2 is a fundamental yet currently unresolved question. Like others, we observed a strong correlation between Spike IgG levels and neutralization capacity (*19, 20*). Although reports on neutralization prevalence and average titers vary widely depending on sampling and detection assay strategies (*21, 22*), our results expand on previous findings by comparing neutralization levels with antigen-specific response over a longer follow-up period with more timepoints than most previous studies. The decline in IgG titers and neutralization often stabilized at different levels later into convalescence, addressing whether decreasing IgG levels eventually plateau. Especially in LIFE whose samples were collected later post-infection and with a longer follow-up period than ADAPT. Spike IgM levels decreased more rapidly than IgG, but were still detectable up to 205 days after diagnosis, much later than previously reported (*1, 19*), and consistent with mathematical modelling of decline of IgM titers in a smaller convalescent cohort (*23*). While our results reveal widely different magnitudes of initial responses and a decrease in neutralizing antibodies titres, most patients have detectable Spike IgG and neutralizing responses more than 5 months after diagnosis, suggesting extended humoral protection, even in those with mild manifestations of the disease.

IgG and IgM against conformational Spike antibody assays have been seldom used, and Spike IgM detection has been challenging. Although many serological assays have reported 100% sensitivity at ∼15 days post-infection (*24*), prevalence studies, vaccine efficacy, and assessment for convalescent COVID-19 plasma donors may not recruit so early post-infection or -vaccination. In this context, and future seroprevalence studies, more sensitive antibody assays will be essential. Flow cytometry assays are used in clinical diagnostics, mainly in the sensitive and specific detection of neuroimmunological autoantibodies in which antigen conformation and discrimination of seropositive patients from healthy controls are critical (*14, 15*). Within the follow up time, the detection of Nucleocapsid and Spike IgG by high capacity commercial assays was significantly less sensitive compared to the flow cytometry assay. Integration of the flow cytometry assay to detect Spike IgG would be valuable to include in the diagnostic pipeline in addition to resource-intensive whole virus neutralization. Given the sensitivity of the flow cytometry assay, this methodology would be ideally suited towards seroprevalence in populations to reveal the true rates of community infection.

The majority of individuals in both cohorts were treated in the community. COVID-19 severity, from mild to hospitalization, was associated with an antibody immune response against SARS-CoV-2 that was reactive toward an increasing number of SARS-CoV-2 antigens, as recently reported (*25*). As our cohorts included only convalescent individuals, the role of broad polyantigenic immunoreactivity in the acute response of hospitalized patients remains unknown. Indeed, reports of patients with absent humoral immune responses have hinted at the role of T cells and innate immune response during the acute disease. Nonetheless, the presence of a broad polyantigenic viral immunoreactivity can be useful to monitor the quality of the antibody response after vaccination.

Whilst the correlation of Spike IgG levels with viral neutralization was strong, high Spike IgM levels were also associated with high viral neutralization in some, especially during the early convalescent days. A lack of somatic mutations was observed in hundreds of cloned neutralizing human antibodies from convalescent patients (*26*). In addition, many antibody precursor sequences were observed in naïve B cells from pre-pandemic patient samples, highlighting the importance of pre-existing germline antibody sequences in the neutralization response. The lack of somatic mutations observed in IgG may be consistent with IgM being potent in a neutralization response as both isotypes could have similar affinity binding sites for Spike, but with multiple binding sites per molecule on IgM, the avidity for Spike would be higher.

Full virus neutralization and prevention of virus-cell fusion were associated. Whilst many assays aim to assess neutralization surrogates outside of level 3 biosafety laboratories, key differences were observed between Spike-driven virus-cell fusion and the authentic SARS-CoV-2 assay. In our study, the particle to transduction ratio in the virus-cell fusion assay was much higher than the SARS-CoV-2 neutralization assay. This is consistent with the respective infectivity of HIV-1 compared to respiratory viruses such as SARS, non-SARS coronaviruses, and influenza (*17, 18*). The virus-cell fusion assay involves a single round of infection, whereas the full virus in the neutralization assay is replication-competent and undergoes multiple rounds of replication over a three day culture. Therefore, the spread of the virus must be considered alongside the capacity of antibodies to inhibit the initial single particle entry and blocking of the virus spread between cells. Although the pseudotyping fusion assay had lower sensitivity, most individuals across both cohorts had titers in this assay with potency ranking similar to full virus neutralization.

Transfusion of convalescent COVID-19 plasma has been proposed as a therapy, with >70 ongoing randomized controlled trials. The few clinical trials to date have supported an acceptable safety profile, but evidence regarding efficacy is mixed (*27*). Most unsuccessful trials included donors with unknown neutralizing status or FDA-classified low titers, whereas successful trials either used high-titer or convalescent COVID-19 plasma delivered within three days of hospitalization (*28*). Indeed, utility of convalescent plasma is improved by donations during early disease stages and by selecting donors with high neutralization antibody titers (*29-31*). Our findings that the immunological response to SARS-CoV-2 is widely heterogeneous, with large variations in SARS-CoV-2 antibodies and neutralization, polyantigenic immunoreactivity, and longitudinal responses complement these assertions. To take into account the first phase of decay observed during early convalescence, we propose an optimal window for plasmapheresis, up to 100 days post-diagnosis. Furthermore, the occurrence of a small group of individuals, termed “high and elite responders” with highly neutralising, broad, and sustained SARS-CoV-2 antibody responses over time, may be due to the rapid and lasting generation of memory B cells (*32*). These patients were likely to be hospitalized older males. Alongside appropriate serology screening programs, the targeted recruitment for plasma donations could help to identify optimal convalescent resources available within affected communities.

A clear advantage of the methodologies used in this study is the capacity of both level 2 biosafety pseudotyped fusion and flow cytometry assays to monitor the effects of viral polymorphisms in real time. Indeed with acceleration of global viral spread, we are now observing evolution of viral fitness and/or immune escape across millions of infected people. The SARS-CoV-2 fitness gain of D614G (*9*) appeared very early in the pandemic and still represents the majority of viral infections globally (>80%)(*10*). Zoonosis of a virus is often followed by finer tuning of replication, as observed in the 2014 Ebola outbreak, in which the variant A82V enabled more efficient receptor NPC1 usage (*33*). Although D614G is a polymorphism outside of the RBD, it significantly impacts the RBD positioning and Spike quaternary structure. The release of hydrogen bonds leading to structural changes are proposed to expose Spike to increase ACE2-dependent fusion (*34*). RBD exposure in the D614G variant may explain the association with great inhibition of virus-cell fusion in patients who recognized the D614G Spike variant. These results are consistent with recent studies in hamsters (*9*) and preliminary data on protection from the ongoing vaccine human trials in areas where the D614G Spike variant remains prevalent. However, our data also highlighted that a subgroup of patients who displayed limited antibody binding to D614G Spike also had reduced virus neutralization irrespective of the viral variant that had infected them. This could be a major concern for vaccine candidate design, especially given the emergence of the S477N/D614G polymorphism in the majority of patients that were infected in the Australian second wave and in Europe (*12, 13*). Seroconversion was observed in all patients from the first and second wave, and good antibody binding to Wuhan-1 D614 and D614G, but there also was a significant decrease in binding and fusion inhibition to S477N/D614G Spike independent of the variant that had infected individuals. Therefore the emergence of the additional polymorphism S477N/D614G could represent an immune evasive variant leading to less antibody immunoreactivity and a resistance to virus neutralization, which could imply a need for periodic variation in vaccine design, as for the influenza vaccine (*35*). Indeed, the mapping of Spike monoclonal antibody escape *in vitro* has recently shown that S477N/D614G is broadly resistant to many neutralizing antibody clones (*36*). Whilst the mechanism behind these observations is unknown, the appearance of a N-glycosylation site within Spike RBD could lead to glycan shielding, as in HIV (*37*), and our evidence that S477N/D614G-infected patients have a similar binding to this variant, albeit reduced, compared to first wave patients, may suggest changing the Wuhan-1 D614 Spike to the S477N/D614G variant in vaccine generation may not overcome the resistance of this variant to the neutralising antibody response. As antibodies against Spike harness the majority of neutralizing activity, selecting the optimal Spike variants in monovalent or multivalent vaccine strategies may be critical.

Our study has important translatable implications to understand the natural history of COVID-19, and post-infection and vaccination-induced immunity. We have highlighted that molecular epidemiology and sero-surveillance will both be required to detect emerging polymorphisms. Furthermore, sensitive monitoring of antibody binding and neutralization capacity will be paramount in vaccine design strategy and convalescent plasma therapy, and in seroprevalence studies, and this would require involvement of more rapidly adaptive methodologies to characterize the magnitude of the neutralization antibody responses against emerging variants.

## Materials and Methods

### Subjects

This study investigated two cohorts of RT-PCR-confirmed convalescent individuals recruited from February to October 2020 in Australia (Table 1 and Fig. 1A). The Adapting to Pandemic Threats (ADAPT) cohort included 83 patients diagnosed at a community-based fever clinic whose sera was collected at two time points post PCR-positivity during the first wave (March-August, n=166 samples). The second wave included sera from 17 patients recruited between July to October. The Australian Red Cross Lifeblood (Lifeblood) cohort (LIFE) included 645 sera samples from 159 donors collected at multiple timepoints post-PCR-positivity (at least 28 days post-recovery) from volunteers presenting to Lifeblood for whole blood or plasma donation. The disease severity of ADAPT patients ranged from mildly symptomatic (mild), community-managed (moderate) to critically unwell and hospitalized (hosp), whereas the self-reported disease severity of LIFE donors included community-managed (non-hosp) and hospitalized (hosp) (Table 1). A healthy adult non-infected pre-pandemic cohort was collected in Australia and consisted of healthy and non-inflammatory neurological disorder donors (n=24). No re-exposure to SARS-CoV-2 and no re-infection was reported. Ethics approval for this study was granted by St Vincent’s Hospital (2020/ETH00964) and Lifeblood (30042020) Research Ethics Committees. Written consent was obtained from all ADAPT patients. In LIFE, the donor consent form included a statement that blood donation may be used in research.

### Flow cytometry cell-based assay for detection of SARS-CoV-2 antibodies

A flow cytometry cell-based assay detected patient serum antibodies against SARS-CoV-2 antigens as for neuroimmunological autoantibodies (*14, 15*). SARS-CoV-2 full-length Spike (Wuhan-1 D614, V367F, G476S, V483A, D614G, S477N/D614G, and S477N/D614G/V1068F) (*10, 11*), Membrane, and Envelope proteins were expressed on transfected HEK293 cells. Serum (1:80) was added to live Spike-expressing cells, and Membrane-, and Envelope-expressing cells were treated with 4% paraformaldehyde and 0.2% saponin, followed by AlexaFluor 647-conjugated anti-human IgG (H+L) (ThermoFisher Scientific) or anti-human IgM (A21249, ThermoFisher Scientific). Cells were acquired on the LSRII flow cytometer (BD Biosciences). Patients were SARS-CoV-2 antibody-positive if their delta median fluorescence intensity (ΔMFI L=L MFI transfected cells – MFI untransfected cells) was above the positive threshold (mean ΔMFI+4SD of 24 pre-pandemic controls) in at least two of three quality-controlled experiments (*14*). Binding to Spike variants was expressed as a percentage of reduced binding compared to Spike. Data was analysed using FlowJo 10.4.1 (TreeStar, USA), Excel (Microsoft, USA), and GraphPad Prism (GraphPad Software, USA).

### Commercial SARS-CoV-2 ELISA

Nucleocapsid IgG assay on the ARCHITECT-I (Abbott Diagnostics, USA), quantitative Spike-1/Spike-2 (S1/S2) IgG on LIASON-155 XL (DiaSorin S.p.A, Italy), and Spike (S1) IgG immunoassay (EUROIMMUN, Germany) were performed. Samples were reported positive if the signal was greater than the published cut-off value (>1.4). Signal to cut-off ratios were used.

### SARS-CoV-2 viral-cell fusion assay

The hACE2 ORF (Addgene# 1786) was cloned into a 3rd generation lentiviral expression vector and clonal stable ACE2-expressing Hek293T cells were generated by lentiviral transductions (*38*). Lentiviral particles pseudotyped with SARS-CoV-2 Spike envelope were produced by co-transfecting Hek293T cells with a GFP encoding lentiviral plasmid HRSIN-CSGW (*39*), psPAX2, and plasmid expressing C-terminal truncated Spike (pCG1-SARS-2-S Delta18) (*40*) including D614 or D614G (*38*). Neutralization activity of sera was measured using a single round infection of ACE2-HEK293T with Spike-pseudotyped lentiviral particles. Virus particles were incubated with serially diluted donor sera for 1 hour at 37°C. Virus-serum mix was then added onto ACE2-HEK293T cells (2.5×10^3^/well) in a 384-well plate. Following spinoculation at 1200g for 1 hour at 18°C, the cells were moved to 37°C for 72 hours. Entry of Spike particles was imaged by GFP-positive cells (InCell Analyzer) followed by enumeration with InCarta software (Cytiva, USA). Neutralization was measured by reduction in GFP expression relative to control group infected with the virus particles without any serum treatment.

The virus entry pathway in VeroE6, used in live virus neutralization assays, is primarily endosomal (*41*). In contrast, cells derived from nasopharyngeal tissues, express ACE2 in addition to the surface serine protease TMPRSS2 which drives virus-cell membrane fusion and can signficantly enhance viral entry (*40*). To address viral neutralization in the presence of ACE2 and TMPRSS2, a portfolio of Hek293T expressing clonal cell lines with ACE2 and TMPRSS2 (HekAT) was generated. The coexpression of ACE2 and TMPRSS2 led to a series of increasingly permissive cell lines that were readily susceptible to SARS-CoV-2 cytopathic effects, VeroE6, HekAT14, HekAT10, HekAT24 (Fig. S3).

### High content fluorescdent live SARS-CoV-2 neutralization assay

Sera were serially diluted and mixed in duplicate with an equal volume of virus solution at 1.5×10^3^ TCID50/mL. After 1 hour of virus-serum coincubation at 37°C, 40μL were added to equal volume of freshly-trypsinized VeroE6 cells, and three clonal HekAT cells in 384-well plates (5×10^3^/well) selected on SARS-CoV-2 permissiveness. After 72h, cells were stained with NucBlue (Invitrogen, USA) and the entire well was imaged with InCell Analyzer. Nuclei counts, proxy for resulting cytopathic effect, were compared between convalescent sera, mock controls (defined as 100% neutralisation), and infected controls (defined as 0% neutralization) using the formula; % viral neutralization = (D-(1-Q))x100/D, where Q = nuclei count normalized to mock controls, and D = 1-Q for average of infection controls (InCarta software).

### Enumeration of SARS-CoV-2 particles

Live SARS-CoV-2 and lentiviral particles were stained using SYTO™ RNASelect™ Green Fluorescent cell Stain (Invitrogen, USA) at a final concentration of 10µM for 30 minutes at 37°C in freshly thawed unpurified viral particles. Particles were then diluted 1/10 and 1/100 in sterile PBS and then adhered to Poly-L-Lysine coated glass bottom 96-well Greiner Sensoplates (Sigma Aldrich, USA) through spinoculation at 1200g for 1 hour at 18°C. Particles were either imaged live or immune-fluorescently counter-stained using a rabbit polyclonal SARS-CoV-2 Nucleocapsid antibody, followed by Alexa647-conjugated goat anti-rabbit IgG (Novus Biologicals, USA). Viral particles were then imaged and quantified as previously described (*42*). Particle to infectivity ratios were determined by dividing the total particle count per ml with the calculated TCID50/ml. Particle to GFP transduction ratios were used for lentiviruses.

### SARS-CoV-2 Spike sequencing and analysis

Clinical respiratory samples were sequenced using an existing amplicon-based Illumina sequencing approach. The raw sequence data were subjected to an in-house quality control procedure before further analysis as reported in (*43*). Non-synonymous SARS-CoV-2 Spike mutations (read frequency >0.8, minimum coverage 10x) were inferred from variant calling files during bioinformatic analysis using phylogenetic assignment of named global outbreak lineages (PANGOLIN)(*11*). All consensus SARS-CoV-2 genomes identified have been uploaded to GISAID (www.gisaid.org).

### Statistics

Statistical analyses were performed in R v4.0.3. Loess curves were generated using ggplot2 v3.3.2. For categorical variables, a log-linear model was fitted and Pearson residuals plotted in a mosaic plot (MASS v7.3-51.6). Shapiro-Wilk test was used to test for normality in continuous variables and a Dwass-Steel-Critchlow-Fligner test was used to test for significance between continuous and categorical variables. Correlations were measured using the Spearman method (psych v2.0.8). Virus-cell fusion and neutralization data were fitted using an exponential decay curve (Origin Lab). Patient curves unable to be fitted, <3 collection dates or low viral fusion and neutralization, were undetermined. Statistical significance was determined as p<0.05.

## Data Availability

Correspondence and requests for data should be addressed to FB and may be limited due to ethical considerations. Plasmids transfer should be obtained through a MTA.

## Acknowledgements

We thank all the patients and donors who participated in this study. We thank Drs. Suat Dervish, Edwin Lau, and Maggie Wang for providing advices at the Flow Cytometry Core Facility of the Westmead Research Hub. We thank Ms Rebecca Rielly and the Operation Team at Kids Research for providing acces to the PC2 facility;

## Author contributions

FB, SGT, AK, and GM designed the study. FT, DP, DV, AOS, AA, VM, FXL, GW, RR, VS, WR, PC conducted and analysed experiments. GD, GM, DRD, IG, VH, DI enrolled and managed patients. FT, AOS, AA, and DRD wrote the manuscript first draft and FT prepared Figs and tables. FB, SGT, AK, and GM designed and coordinated research and verified results. All authors reviewed the draft before submission;

## References

1. D. F. Gudbjartsson et al., Humoral Immune Response to SARS-CoV-2 in Iceland. N Engl J Med 383, 1724–1734 (2020).

2. B. Isho et al., Persistence of serum and saliva antibody responses to SARS-CoV-2 spike antigens in COVID-19 patients. Sci Immunol 5, (2020).

3. F. J. Ibarrondo et al., Rapid Decay of Anti–SARS-CoV-2 Antibodies in Persons with Mild Covid-19. New England Journal of Medicine 383, 1085–1087 (2020).

4. M. Pollán et al., Prevalence of SARS-CoV-2 in Spain (ENE-COVID): a nationwide, population-based seroepidemiological study. Lancet 396, 535–544 (2020).

5. T. F. Rogers et al., Isolation of potent SARS-CoV-2 neutralizing antibodies and protection from disease in a small animal model. Science 369, 956–963 (2020).

6. M. Hoffmann et al., Camostat mesylate inhibits SARS-CoV-2 activation by TMPRSS2-related proteases and its metabolite GBPA exerts antiviral activity. bioRxiv, (2020).

7. A. Wajnberg et al., Robust neutralizing antibodies to SARS-CoV-2 infection persist for months. Science 370, 1227–1230 (2020).

8. Q. Li et al., The Impact of Mutations in SARS-CoV-2 Spike on Viral Infectivity and Antigenicity. Cell 182, 1284–1294 e1289 (2020).

9. Y. J. Hou et al., SARS-CoV-2 D614G variant exhibits efficient replication ex vivo and transmission in vivo. Science, (2020).

10. B. Korber et al., Tracking Changes in SARS-CoV-2 Spike: Evidence that D614G Increases Infectivity of the COVID-19 Virus. Cell 182, 812–827 e819 (2020).

11. Z. Liu et al., Landscape analysis of escape variants identifies SARS-CoV-2 spike mutations that attenuate monoclonal and serum antibody neutralization. bioRxiv, 2020.2011.2006.372037 (2020).

12. A. T. Chen, K. Altschuler, S. H. Zhan, Y. A. Chan, B. E. Deverman, COVID-19 CG: Tracking SARS-CoV-2 mutations by locations and dates of interest. bioRxiv, (2020).

13. E. B. Hodcroft et al., Emergence and spread of a SARS-CoV-2 variant through Europe in the summer of 2020. medRxiv, 2020.2010.2025.20219063 (2020).

14. F. Tea et al., Characterization of the human myelin oligodendrocyte glycoprotein antibody response in demyelination. Acta Neuropathol Commun 7, 145 (2019).

15. F. Graus et al., A clinical approach to diagnosis of autoimmune encephalitis. Lancet Neurol 15, 391–404 (2016).

16. D. R. Burton, L. Hangartner, Broadly Neutralizing Antibodies to HIV and Their Role in Vaccine Design. Annu Rev Immunol 34, 635–659 (2016).

17. P. J. Klasse, Molecular determinants of the ratio of inert to infectious virus particles. Prog Mol Biol Transl Sci 129, 285–326 (2015).

18. K. Martin, A. Helenius, Transport of incoming influenza virus nucleocapsids into the nucleus. J Virol 65, 232–244 (1991).

19. A. S. Iyer et al., Persistence and decay of human antibody responses to the receptor binding domain of SARS-CoV-2 spike protein in COVID-19 patients. Sci Immunol 5, (2020).

20. S. L. Klein et al., Sex, age, and hospitalization drive antibody responses in a COVID- 19 convalescent plasma donor population. J Clin Invest 130, 6141–6150 (2020).

21. D. F. Robbiani et al., Convergent antibody responses to SARS-CoV-2 in convalescent individuals. Nature 584, 437–442 (2020).

22. X. Wang et al., Neutralizing Antibodies Responses to SARS-CoV-2 in COVID-19 Inpatients and Convalescent Patients. Clin Infect Dis, (2020).

23. A. K. Wheatley et al., Evolution of immunity to SARS-CoV-2. medRxiv, 2020.2009.2009.20191205 (2020).

24. A. Bryan et al., Performance Characteristics of the Abbott Architect SARS-CoV-2 IgG Assay and Seroprevalence in Boise, Idaho. J Clin Microbiol 58, (2020).

25. E. Shrock et al., Viral epitope profiling of COVID-19 patients reveals cross-reactivity and correlates of severity. Science 370, eabd4250 (2020).

26. C. Kreer et al., Longitudinal Isolation of Potent Near-Germline SARS-CoV-2- Neutralizing Antibodies from COVID-19 Patients. Cell 182, 843–854 e812 (2020).

27. K. L. Chai et al., Convalescent plasma or hyperimmune immunoglobulin for people with COVID-19: a living systematic review. Cochrane Database Syst Rev 10, Cd013600 (2020).

28. H. Abolghasemi et al., Clinical efficacy of convalescent plasma for treatment of COVID-19 infections: Results of a multicenter clinical study. Transfus Apher Sci 59, 102875 (2020).

29. E. Salazar et al., Significantly Decreased Mortality in a Large Cohort of Coronavirus Disease 2019 (COVID-19) Patients Transfused Early with Convalescent Plasma Containing High-Titer Anti-Severe Acute Respiratory Syndrome Coronavirus 2 (SARS-CoV-2) Spike Protein IgG. Am J Pathol, (2020).

30. M. J. Joyner et al., Convalescent Plasma Antibody Levels and the Risk of Death from Covid-19. N Engl J Med, (2021).

31. R. Libster et al., Early High-Titer Plasma Therapy to Prevent Severe Covid-19 in Older Adults. N Engl J Med, (2021).

32. A. Arunasingam et al., Long-Term Persistence of Neutralizing Memory B Cells in Sars-Cov-2. SSRN, (2020).

33. W. E. Diehl et al., Ebola Virus Glycoprotein with Increased Infectivity Dominated the 2013-2016 Epidemic. Cell 167, 1088–1098 e1086 (2016).

34. L. Yurkovetskiy et al., Structural and Functional Analysis of the D614G SARS-CoV- 2 Spike Protein Variant. Cell 183, 739–751 e738 (2020).

35. K. Houser, K. Subbarao, Influenza vaccines: challenges and solutions. Cell Host Microbe 17, 295–300 (2015).

36. L. Liu et al., Potent Neutralizing Antibodies Directed to Multiple Epitopes on SARS- CoV-2 Spike. bioRxiv, 2020.2006.2017.153486 (2020).

37. T. Zhou et al., Quantification of the Impact of the HIV-1-Glycan Shield on Antibody Elicitation. Cell Rep 19, 719–732 (2017).

38. A. Aggarwal et al., Mobilization of HIV spread by diaphanous 2 dependent filopodia in infected dendritic cells. PLoS Pathog 8, e1002762 (2012).

39. M. G. Toscano et al., Efficient lentiviral transduction of Herpesvirus saimiri immortalized T cells as a model for gene therapy in primary immunodeficiencies. Gene Ther 11, 956–961 (2004).

40. M. Hoffmann et al., SARS-CoV-2 Cell Entry Depends on ACE2 and TMPRSS2 and Is Blocked by a Clinically Proven Protease Inhibitor. Cell 181, 271–280 e278 (2020).

41. J. Wei et al., Genome-wide CRISPR Screens Reveal Host Factors Critical for SARS-CoV-2 Infection. Cell, (2020).

42. S. G. Turville, M. Aravantinou, H. Stossel, N. Romani, M. Robbiani, Resolution of de novo HIV production and trafficking in immature dendritic cells. Nat Methods 5, 75–85 (2008).

43. R. J. Rockett et al., Revealing COVID-19 transmission in Australia by SARS-CoV-2 genome sequencing and agent-based modeling. Nat Med 26, 1398–1404 (2020).

