## Supplementary Material_text and Figures for "SARS-CoV-2 neutralizing antibodies; longevity, breadth, and evasion by emerging viral variants"

### Supplementary Figures

**Fig. S1. Correlation matrices for antibody responses and demographic data in ADAPT (A) and LIFE (B).** Continuous measures of demographics (age, days post PCR-positivity) and antibody titers (Nucleocapsid, Spike IgG and IgM, and Membrane), were compared. R-square values are placed in the boxes (top-right) with P value significance shown in *. Bottom-left show correlation plots with Loess line (red).

**Fig. S2. Spike IgG decay profile of high plasmapheresis donors.** Spike IgG titer over time is shown from n=30 LIFE donors who underwent >10 plasmapheresis donations. Despite high donations, donors with high Spike IgG titers decreased but stabilized at mid-levels, whereas donors with mid to low Spike IgG titers, stabilized at low titers. No donors seroreverted nor became negative for Spike IgG.

**Fig. S3. Hyper-permissiveness of Hek cell lines.** HekAT clonal cell lines and VeroE6 were infected with serially diluted SARS-CoV-2 and monitored for cytopathic effect (CPE) at 72 hours post-infection. **(A)** Cell nuclei were stained with NucBlue and CPE quantified as % live cells. HekAT clones showed varying degrees of permissiveness to SARS-CoV-2 infection with HekAT24 being orders of magnitude more susceptible than VeroE6. WT Hek293T cells were refractory to infection and were used as negative control. (**B**) Bright-field images showing CPE in WT Hek and HekAT clonal lines. Images were acquired using InCell high throughput imaging system. Magnification is 10X for all images. Representative images are shown.

**Fig. S4. GFP expression and positive individual status against two new Spike variants.** **(A)** Expression of GFP reporter molecule expressed via the transcription of Spike-GFP monocistron in the pIRES2 plasmids was similar across all analysed variants allowing accurate comparisons between IgG levels between variants. **(B)** Pre-pandemic controls were below the threshold, whereas all first and second wave of ADAPT samples were above the positive threshold (Control + 4SD).

**Fig. S5. Individuals with reduced immunoreactivity to D614G Spike variant were more likely to be females.** Mosaic plots of ADAPT (left) and LIFE (right) show individuals with restricted binding to Spike variant, i.e. reduced binding to D614G, were more likely to be females.

Supplementary Figure 1.

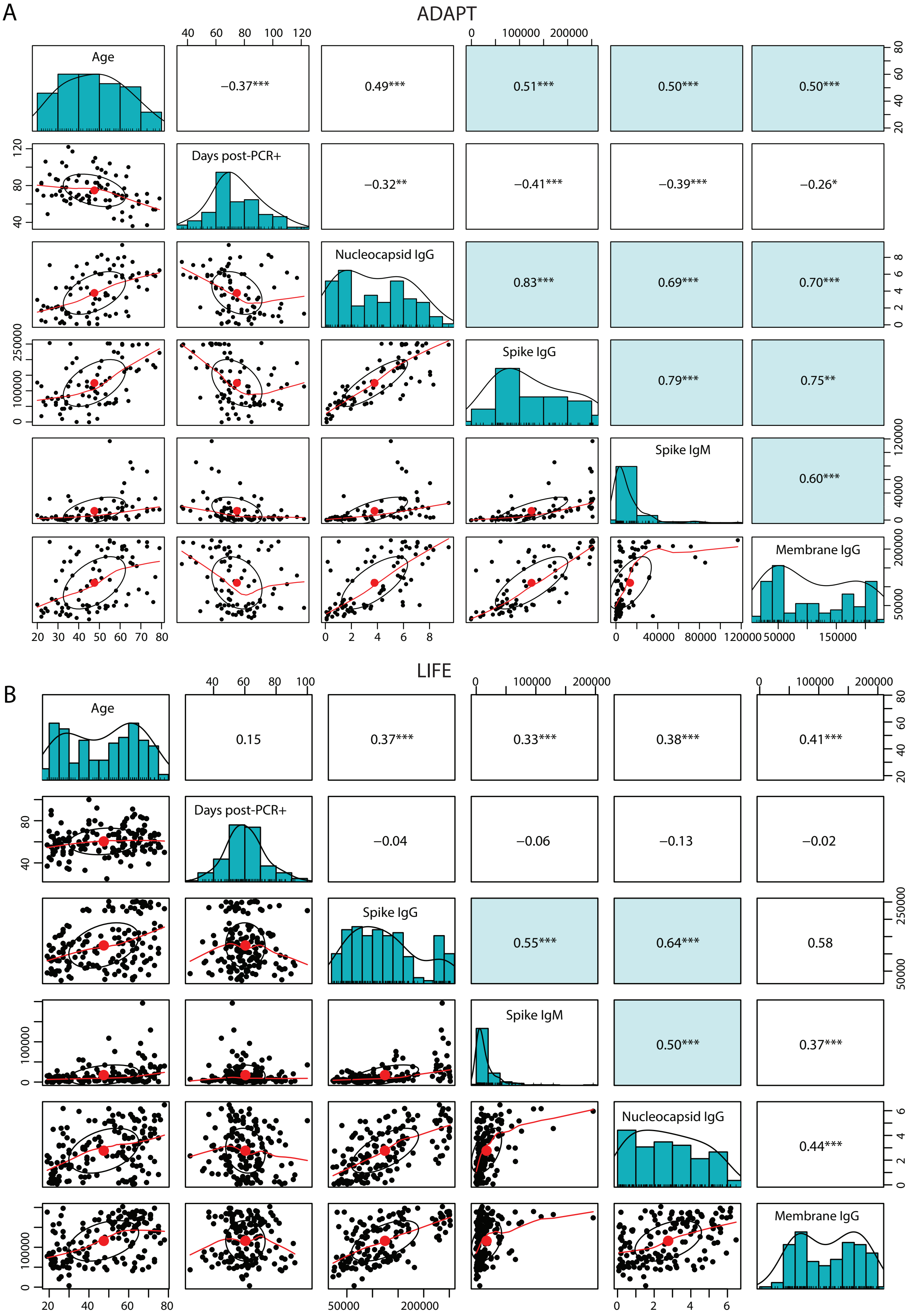

### Supplementary Figure 2.

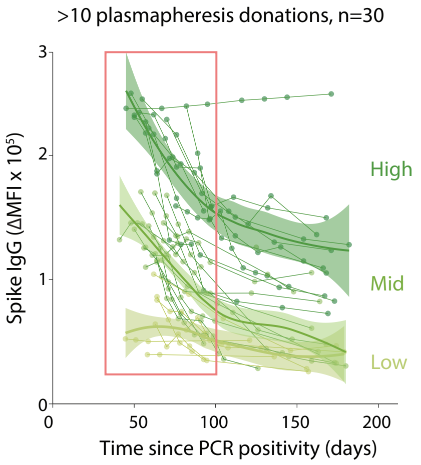

### Supplementary Figure 3

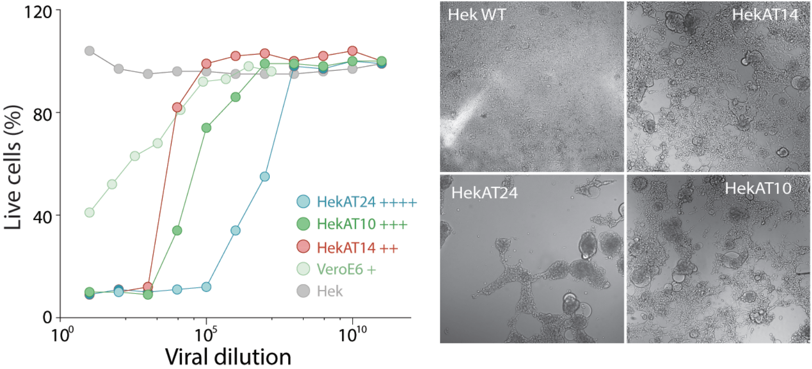

### Supplementary Figure 4

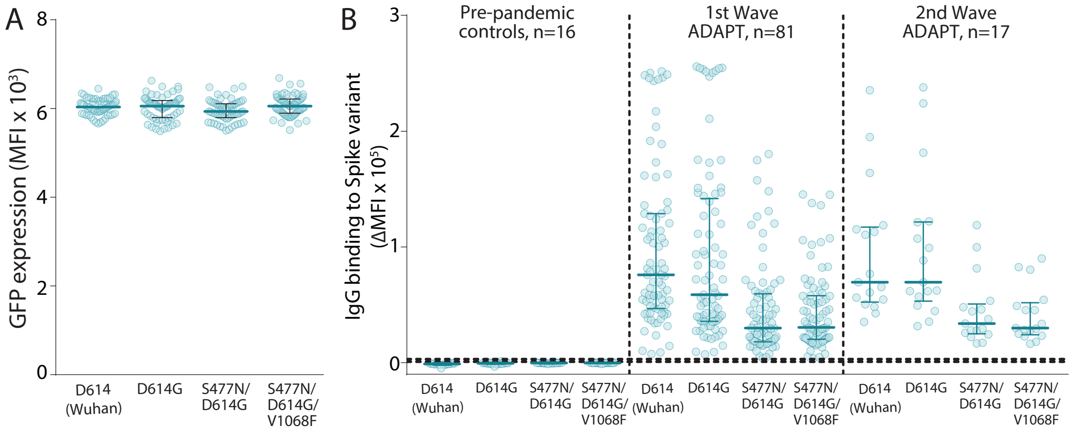

### Supplementary Figure 5

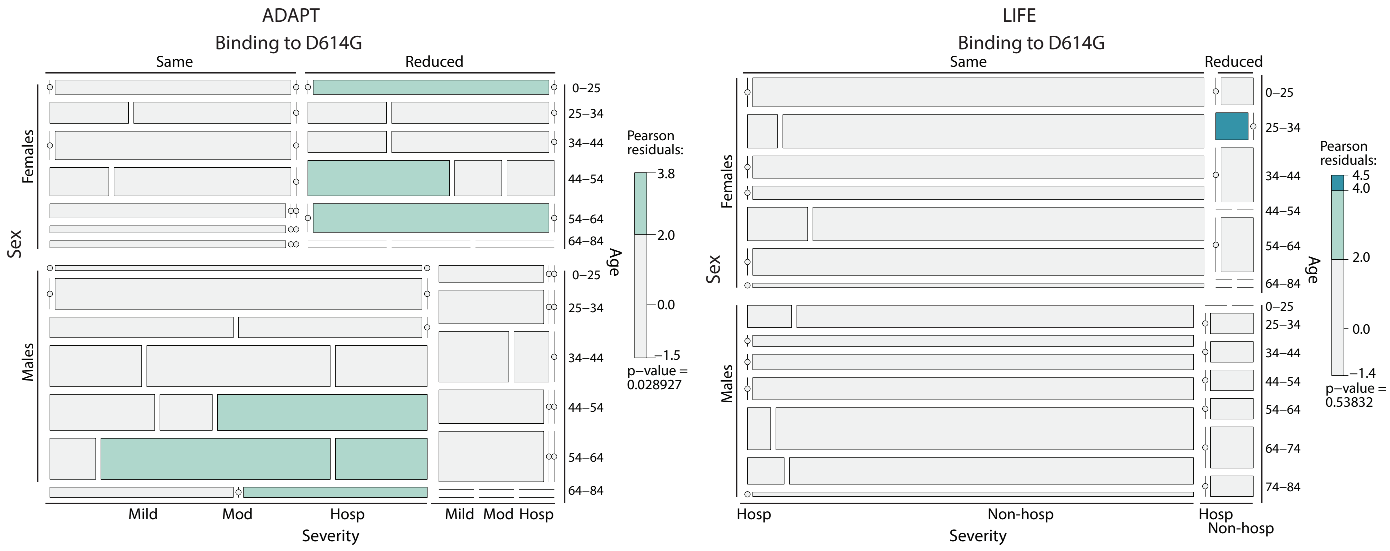
